# Opioidergic pain relief in humans is mediated by beta and high-gamma modulation in limbic regions

**DOI:** 10.1101/2025.03.03.25323046

**Authors:** Jacob C. Garrett, Sierra Wilson, Alexander Jessup, Michael G. Brandel, Caleb S. Nerison, Ahmed M. Raslan, Sharona Ben-Haim, Eric Halgren

**Author notes:** Equal contribution (Sharona Ben-Haim, Eric Halgren).

## Abstract

The neurophysiological mechanisms underlying opioid analgesia remain poorly understood, limiting the development of non-addictive alternatives. Fentanyl and hydromorphone were administered to patients experiencing semi-chronic, clinically relevant pain following surgical implantation of intracranial electrodes for seizure localization. Opioids suppressed beta oscillations in lateral amygdala and ventral and dorsolateral prefrontal cortices, while increasing beta power in the medial amygdala and hippocampus. High-gamma amplitude was suppressed in the insula and lateral amygdala, and increased in the cingulate cortex and hippocampus. The magnitude of beta suppression in the ventral prefrontal cortex and beta enhancement in the medial amygdala and hippocampus, as well as high-gamma suppression in the insula, were positively correlated with pain relief in response to a constant dose. These findings identify electrophysiological events in a limbic network that may mediate opioidergic analgesia via nociceptive gating and reduced affective salience of pain, offering insights into the neural basis of pain relief and potential biomarkers for the development of non-addictive opioid alternatives.

## Introduction

Human pain carries significant affective and interpretive components [34,48,87]; yet relative to peripheral and spinal mechanisms, far less is known about its physiology in higher-order telencephalic substrates. In the present study, we utilized bipolar intracranial recordings to isolate signatures of opioid-mediated pain relief in the human forebrain.

Further characterization of these higher-order neurophysiological signatures of clinically-relevant human pain and pharmacological pain relief may guide the development of improved treatments. To our knowledge, only one prior study has successfully predicted spontaneous pain from LFP measures alone [77], a clear step toward individualized biomarkers for analgesic development. Nevertheless, this model was trained separately for each subject on a limited set of brain regions, restricting its generalizability to broader mechanisms or therapeutic targets [69].

Cross-sectional EEG studies of chronic pain have identified positive associations between frontal beta power and arthritic pain severity [79], as well as enhanced beta coherence in fibromyalgia [4]. Patients with neuropathic pain show increased power in the alpha-beta range [20,82], and a systematic review of EEG and MEG studies have concluded that enhanced beta and theta oscillations are characteristic of chronic pain conditions generally [90].

Opioids remain among the most powerful non-surgical analgesics, yet their neurophysiological effects on human EEG are surprisingly understudied. One scalp EEG study in healthy patients found decreases in beta and alpha power with moderate doses of an oral opioid [11], while a more recent report observed decreases in beta and theta power during methadone therapy [58].

Thus, in the present study, utilizing intracranial recordings from epilepsy patients undergoing seizure monitoring, we analyzed the effects of intravenous opioids on beta (15-25 Hz) amplitude. Beta oscillations are especially prominent in intracranial EEG, particularly in frontal and sensorimotor regions densely sampled here and implicated in chronic pain [13]. We also analyzed the effects of opioids on high gamma (HG; 70-190 Hz) amplitude, a proxy for population firing, based on prior iEEG studies linking HG to acute pain ratings [12].

To our knowledge, this is the first study to identify consistent neural signatures of opioid effects on human local field potentials, as well as the first to report neurophysiological predictors of pain relief in a clinical context.

## Materials and methods

### Patient population

The participants gave written informed consent to participate in this study, and the study was approved by the University of California and Oregon Health and Science University Institutional Review Boards in accordance with the Declaration of Helsinki. Patients were compensated for their participation. Patients were recruited from a population with long-standing drug-resistant complex partial epilepsy undergoing intracranial EEG monitoring with implanted depth electrodes to localize seizure foci. While awaiting spontaneous seizures, the patients were offered the opportunity to consent to research procedures. Regions of the frontal lobe and limbic system frequently implanted for clinical monitoring are also implicated in the affective-interpretive dimensions of pain. As a consequence of the surgical implantation procedure, many patients experience some degree of post-surgical pain, which is variable in both severity and required treatment. A subset of four patients (of 26 with meaningful pain and usable data) received at least 10 doses of intravenous opioids unconfounded by putative sleep or peri-ictal periods (defined below), constituting a rare cohort with sufficient clinical and physiological resolution to support quantitative analysis (see Table 1 and Figure 1). All patients had overall cognitive function in the normal range.

**Figure 1.**
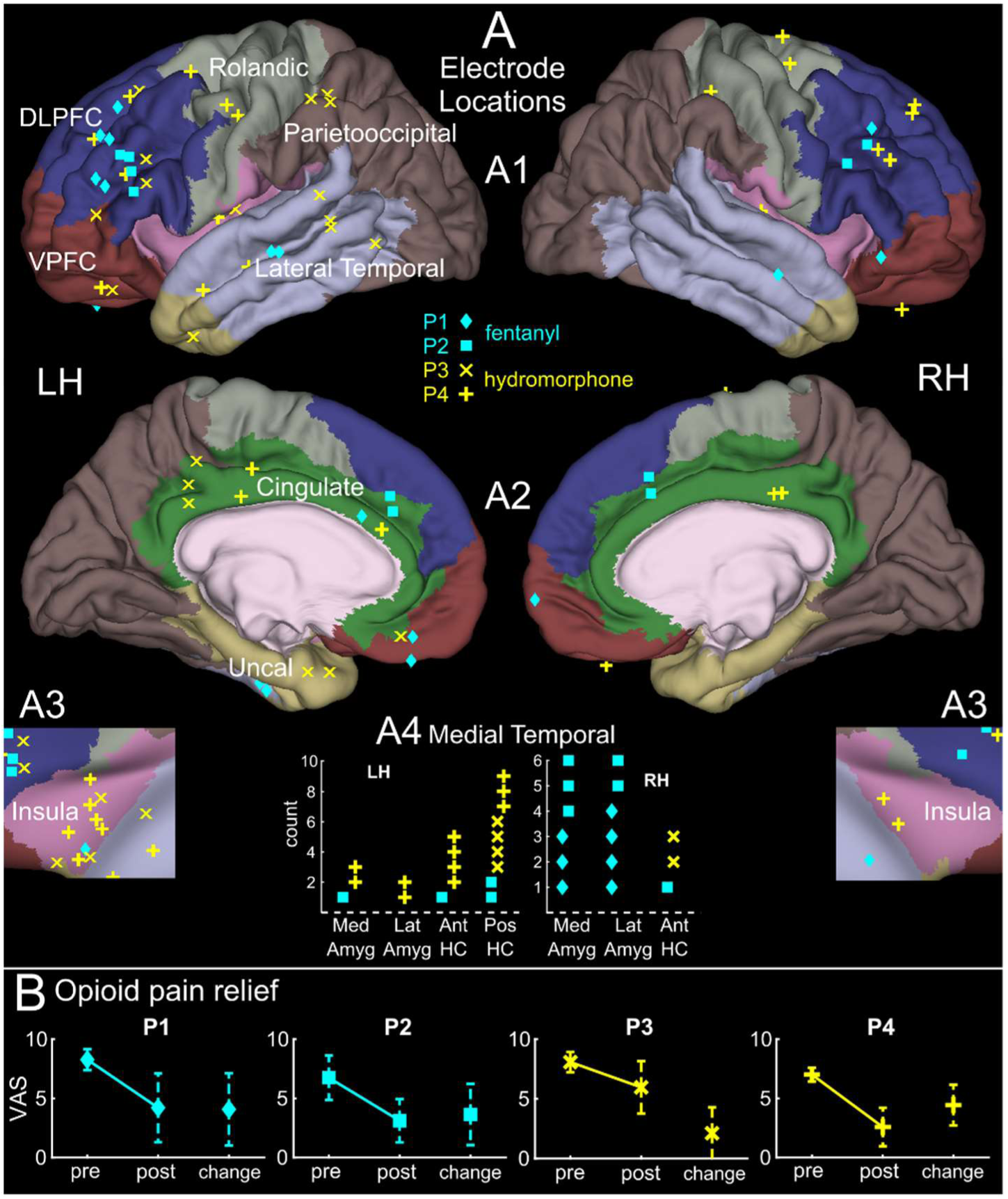
Recording sites and opioid effects on pain. Subjects are denoted by marker shape: diamond (P1), square (P2), X (P3), and cross (P4); drug type is indicated by color (fentanyl[FENT]=cyan, hydromorphone[HMOR]=yellow). **A. Location of recording sites by subject and drug type.** Electrodes on the lateral (A1) and medial (A2) cortical surfaces are shown in anatomically segmented regions (pastel colors), based on superimposed CT and MRI. Insular sites (A3) are made visible by relaxing the curvature of overlying cortex. Medial temporal sites (A4) are shown in a table because they cannot be seen from the surface. **B. Characteristics of pre-drug pain and post-drug pain change**, by subject, mean+SD. Mean VAS decrease pre-post opioid = 3.2 (p<10e-14, paired t-test).

**Table 1.** Subject characteristics. “Drug effect analysis” = number of doses without sleep/seizure interference and with pre-drug pain score. “Pain analysis” = subset with post-drug pain score <90 mins post-administration.

| Subject | Age | Sex | Days | Drug | N Doses<br>(Drug<br>effect) | N Doses<br>(Pain<br>analysis) | Dose<br>(mode) | N<br>Sites | Median<br>(Range)<br>VAS |
| --- | --- | --- | --- | --- | --- | --- | --- | --- | --- |
| P1 | 36-40 | M | 10.7 | Fentanyl | 22 | 14 | 50 mcg | 25 | 5 (0-10) |
| P2 | 21-25 | F | 9.0 | Fentanyl | 33 | 21 | 12.5<br>mcg | 21 | 4 (0-9) |
| P3 | 31-35 | M | 13.0 | Hydro-<br>morphine | 51 | 29 | 1 mg | 29 | 7 (0-10) |
| P4 | 31-35 | M | 15.0 | Hydro-<br>morphine | 14 | 8 | 0.5 mg | 41 | 5.5 (0-10) |

### Data preprocessing

Archival SEEG recordings were processed and reformatted using standard methods. A notch filter was applied at 60 Hz and its harmonics up to the Nyquist frequency. Data were then resampled to 1000 Hz from native sampling rates (500 or 1024 Hz) using a delay-compensated FIR antialiasing low-pass filter. To isolate local signal, monopolar recordings were re-referenced in bipolar montage using adjacent contacts (5 mm pitch). Bipolar pairs were excluded if they showed low amplitude (e.g. <30 µV slow-wave during NREM, indicating white matter or CSF), abnormal LFP (e.g. atypical oscillations), frequent interictal discharges (>3/min in medial temporal lobe, >1/min in cortex), or were part of a seizure focus. Time series analysis was performed using custom MATLAB [83] code and FieldTrip [65,83].

Cortical bipolar pairs were selected from non-overlapping contacts. In the hippocampus and amygdala, some overlap was allowed due to limited high-quality channels. Electrode locations were determined by MRI/CT co-registration, followed by automated surface reconstruction by FreeSurfer [26,27] and parcellation utilizing the HCP-MMP1 atlas [32]. In one patient (P3) lacking complete imaging for surface reconstruction, locations were hand-annotated using trajectories drawn from clinical neurosurgery software (ROSA). HCP-MMP1 parcels were aggregated to form regions of interest (ROIs) for cross-subject analysis and localization of opioid effects (Figure 1, Table S4). Medial temporal electrodes were manually localized in co-registered MRI/CT using 3D Slicer [43].

### Epoch selection and artifact rejection

Interictal epileptiform discharges (IEDs) were detected as described previously [21,22]. A high frequency score was computed by smoothing the 70-190 Hz analytic amplitude with a 20 ms boxcar, and a spike-similarity score by cross-covariance with a canonical IED waveform. Both scores were normalized by channel median. Weighted by 13 (HF) and 25 (spike-similarity), IEDs were flagged when the weighted sum exceeded 130. Detected IEDs and intervening epochs were visually reviewed in each channel to confirm detection sensitivity and specificity. Each flagged period, the 300 ms preceding it, and the 700 ms following it were excluded. Putative seizures were defined as dense, sudden clusters of epileptiform detections as above, cross-checked with clinical records, and visually verified in the LFP. Seizure epochs, the 5 minutes preceding, and a period of 2 hours following were excluded to account for spectral disturbances (e.g. HG suppression, slow-wave enhancement) persisting postictally. Epochs with large artifacts, signal interruptions, or unstable spectral features (assessed by combined visual examination of beta, HG, slow-wave, 8-12 Hz alpha, and 4-8 Hz theta amplitudes) suggestive of prolonged or unstable postictal dynamics, epileptiform activity, or residual anesthesia effects were similarly excluded.

### Sleep rejection

Sleep rejection followed previously described methods [22], based on N2/N3 criteria defined by Silber et al. [78]. A slow-wave sleep score was computed as the median slow-wave (0.5-2 Hz) amplitude across channels. Sleep cycles were identified as periods where this score exceeded 3-fold the local 36-hour median, computed thusly to account for amplitude drift. A 25 minute buffer before and after each N2/N3 epoch was excluded to eliminate N1 and REM.

### Testing of drug effect

Administration times of intravenous opioid medications were recorded with minute-scale precision via clinical records. We compared neurophysiological metrics during the period 30-10 minutes preceding opioid administration to the period 10-30 minutes following, which is within the period of peak analgesic effect for intravenous fentanyl and hydromorphone [15,17]. As both are primarily µ-opioid receptor agonists [24] with comparable analgesic efficacy at clinical doses [2,67], we combined them in primary analyses while including a categorical covariate for drug type in mixed-effects models. A potential extended effect of hydromorphone, due to the slower redistribution relative to fentanyl, was tested post-hoc [56,71]. All included opioid doses were separated by at least 60 minutes (see Fig. S1 for distribution of relative administration times and dosages).

### Spectral perturbation plots

For event-related spectral perturbation (ERSP; Fig 2A,B) and spectral shift (Fig. 2C,D) plots, spectral power was computed using a multi-taper time-frequency method (Chronux toolbox [14]). Power spectra were computed for each minute of data, excluding segments rejected for IEDs or artifacts. Each minute was subdivided into continuous, non-overlapping segments of 8 seconds, with the mean spectrum computed across all valid segments. Spectra were log-resampled in the frequency domain using a zero-phase FIR antialiasing filter. Drug-evoked changes were normalized to the pre-drug baseline (30-10 minutes before dose).

**Figure 2.**
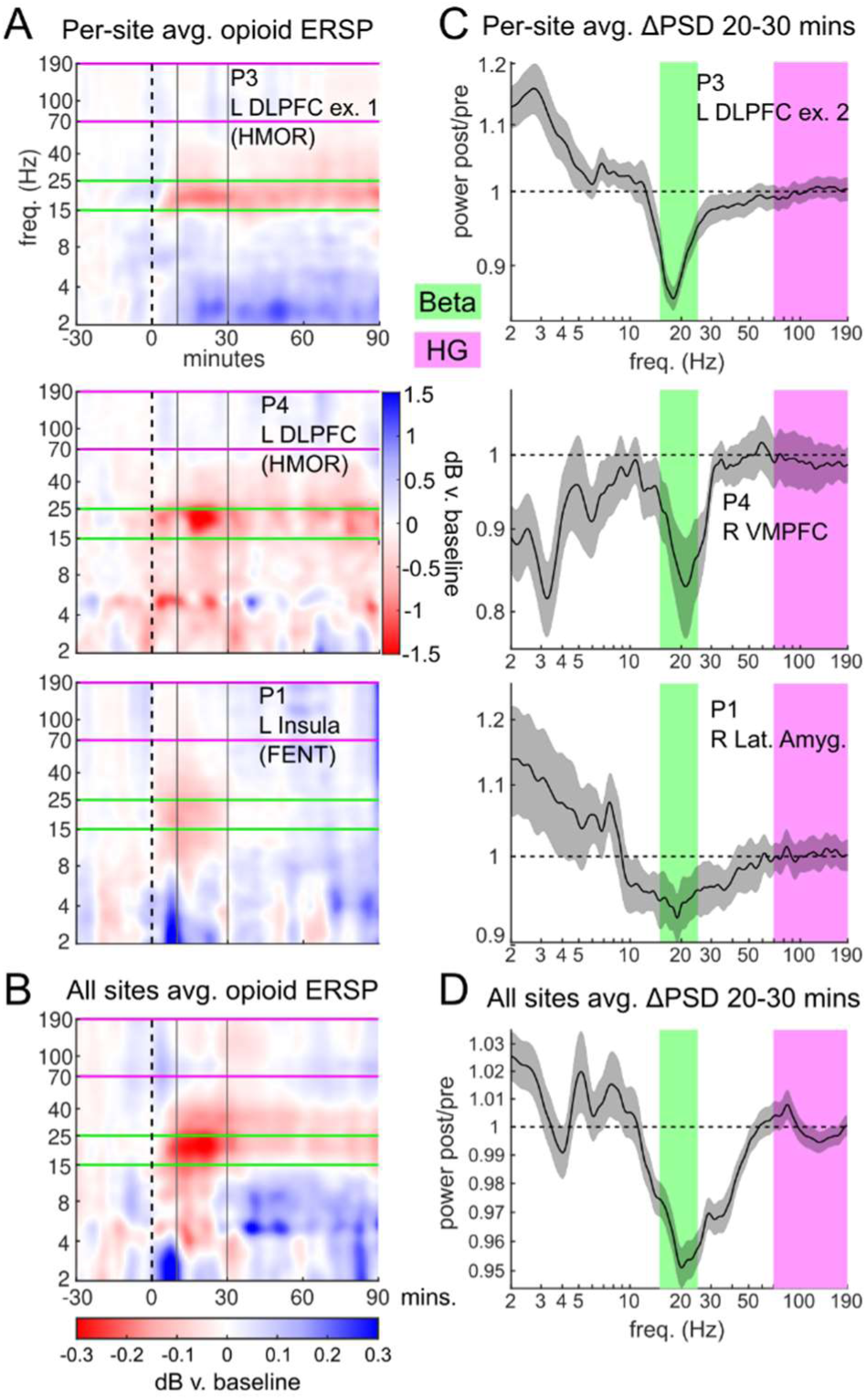
Opioid spectral response. A. Time-frequency spectral response (2 to 190Hz) of single sites to IV opioid injections. Beta (15-25Hz, between green horizontal lines) is suppressed from ∼10-30 min post-injection (black solid lines). Each frequency is normalized to the -30 to -10 minute baseline. **B**. As in A, but the average over all sites in all subjects. **C.** Post-drug (10-30 min) power spectra normalized to pre-drug baseline, for example sites. Beta suppression (green) is evident. **D**. As in C, but the average over all sites in all patients. Gray shaded area indicates standard error of cross-channel mean. All shown sites exhibited significant (p<0.05 FDR) beta suppression during the 10 to 30 minute post-drug period.

In ERSP plots (Fig. 2A,B), the spectral response at each minute post-drug was expressed in dB and divided by pre-drug baseline power. For spectral shift plots (Fig. 2C,D), the average spectral power from 10-30 minutes post-drug divided by the pre-drug baseline to yield the proportional drug-evoked change in LFP power by frequency. For display, results were smoothed over time (ERSP) and frequency (ERSP and change plots) using a Gaussian kernel.

### Evoked amplitude-modulation plots

To examine opioid-evoked changes in beta and high-gamma (HG) amplitude, a third-order Butterworth bandpass filter was applied (15-25 Hz for beta; 70-190 Hz for HG), and the exact Hilbert analytic amplitude computed [50]. To account for long-term spectral drift, amplitude traces were z-scored using the mean and standard deviation of the baseline period for the same trial (30-10 minutes pre-dose). Trials were excluded if >75% of the baseline or >25% of the effect period was rejected (per criteria in **Epoch selection and artifact rejection** and **Sleep rejection)**. Channels were grouped within predefined regions of interest (ROIs) based on anatomical sampling, prior pain literature, and standard neuroanatomical boundaries (Table S4). The average z-scored amplitude 10-30 minutes post-drug was computed per channel for cross-site analyses.

### Site-wise pain-relief correlation estimates

To evaluate neural correlates of pain relief, we computed the correlation between pain relief (VAS score change) pre-to-post opioid and the corresponding change in beta or HG amplitude. Neural measures were extracted from the 30-10 minute pre-drug period and the 10 minutes immediately preceding the post-drug VAS score. Only trials involving the modal opioid dose per subject were included. Trials were excluded if the pre-drug VAS score was >20 minutes before the dose, or if the post-drug VAS score was <20 or >90 minutes after opioid administration. As above, neural amplitude measures were z-scored individually according to the mean and standard deviation of the baseline period of the same trial. Pearson correlation coefficient (R) between amplitude change and VAS change was computed per-site and used in cross-site modeling.

### Cross-site drug-effects and pain relief models

We fit linear mixed-effects (LME) models to assess overall and regional patterns in opioid effects and pain relief, separately for each frequency band (beta/HG) and outcome type (drug effect or pain correlation). Models were fit via maximum likelihood using MATLAB R2019b’s fitlme function [83]. Models predicted z-score of drug effect or R-value of pain-amplitude correlation *y* for each contact *c* in subject *s*:

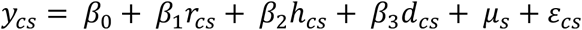

where *r* represents brain region, *h* hemisphere, *d* drug type (HMOR>FENT), and *µ* a random effect on intercept by subject. In all four primary models, random effects collapsed to zero, suggesting minimal unexplained subject-level variance.

To account for clinical context variability, we additionally modeled trial-wise responses using both trial (*t*) and subject as random effects on the intercept.

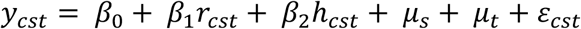

Finally, to test whether opioid-evoked changes predicted pain relief, we modeled site-wise R-values (pain correlation) as a function of drug-effect z-scores, including a random intercept by subject.

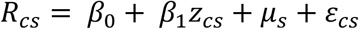

### Control analysis: non-opioid nurse check-ins

To rule out neurophysiological changes due to clinical interaction per se, we applied the same analysis to non-opioid nurse check-ins: timepoints at which the nurse similarly interacted with the patient and recorded a VAS score, but no opioid was administered. These events were only included if no opioid had been given in the preceding 90 minutes and none was administered in the following 30 minutes. While other medications (e.g. acetaminophen) may have been given, no oral or intravenous opioids were administered. We then applied the same LME model used for opioid analysis to assess whether beta or HG amplitude changed during these check-ins, including any region- or hemisphere-specific effects.

## Results

### Patient cohort and pain response

Four patients with >10 analyzable intravenous opioid doses were included. Depth electrodes sampled widespread cortical and limbic regions, with especially dense coverage in prefrontal and medial temporal structures, and substantial sampling of insular and cingulate cortex— though cingulate coverage was anatomically heterogeneous (Fig. 1A, Table S5). All patients reported postoperative pain due to penetration of the scalp and dura by implanted electrodes and were given opioid medication (Table 1). Intravenous opioid doses were followed by a substantial reduction in pain score (VAS rating) in all subjects (Fig. 1B, mean VAS decrease pre-post opioid = 3.2; p<10e-14, paired t-test). Based on prior literature, we hypothesized that opioids would produce reliable suppression of beta-frequency activity in frontal and limbic sites.

### Spectral characterization of opioid-evoked responses

We examined spectral dynamics surrounding opioid administration using bipolar recordings. Concordant with prior literature, a distinct suppression of power centered on the beta frequency range (15-25 Hz) was consistently observed shortly following drug administration in both individual electrodes (Fig. 2A) and across subjects (Fig. 2B). The period 10-30 minutes post-opioid was chosen to measure drug effect, as this falls within the period of peak pain relief for both drugs[15,17]. Power spectral density analysis confirmed the characteristic beta suppression at both single-channel (Fig. 2C) and population levels (Fig. 2D).

### Hydromorphone produces longer-lasting beta suppression

Consistent with the slower redistribution kinetics of hydromorphone (HMOR), beta suppression evoked by HMOR injection was more sustained than that following fentanyl (FENT). We identified sites showing significant beta suppression from 10-30 minutes post-drug (FDR corrected p<0.05) and compared the persistence of this suppression in the 30-50 minute window between HMOR and FENT. Of the 23 sites beta-suppressed by FENT from 10-30 minutes, only 2 (8.7%) remained suppressed from 30-50 minutes. In contrast, of the 31 HMOR sites suppressed in the earlier period, 14 (42.2%) remained suppressed during the same period (χ^2^ test, p=0.004; Table 2).

**Table 2.** Hydromorphone evokes longer beta suppression than fentanyl. “Active”=site-wise effects FDR corrected p<0.05. Only sites active at 10-30 mins were tested at 30-50 mins. Highlighted: χ^2^ test by drug and duration p=0.004.

| Drug | N sites | Active 10-30 | Active 30-50 |
| --- | --- | --- | --- |
| Fentanyl | 55 | 23 | 2 |
| Hydromorphone | 65 | 31 | 14 |

### Beta suppression localizes to prefrontal cortex

Beta amplitude (15-25 Hz analytic) was robustly suppressed by IV opioid administration in all subjects, demonstrated by within-subject averages (Fig. 3A) and at the single-channel level (Fig. 3B). Spatial mapping of site-wise responses showed a gradient in which more frontal sites exhibit greater beta suppression—this was evident in subjects P3 and P4, while subjects P1 and P2 had limited posterior sampling (Fig. 3C). Beta suppression was observed bilaterally and was similar between FENT and HMOR (Fig. 3D). A linear mixed-effects model confirmed a significant main effect of opioids on beta suppression, with significant further suppression in ventral prefrontal cortex (VPFC) and dorsolateral prefrontal cortex (DLPFC) (Table 3).

**Figure 3.**
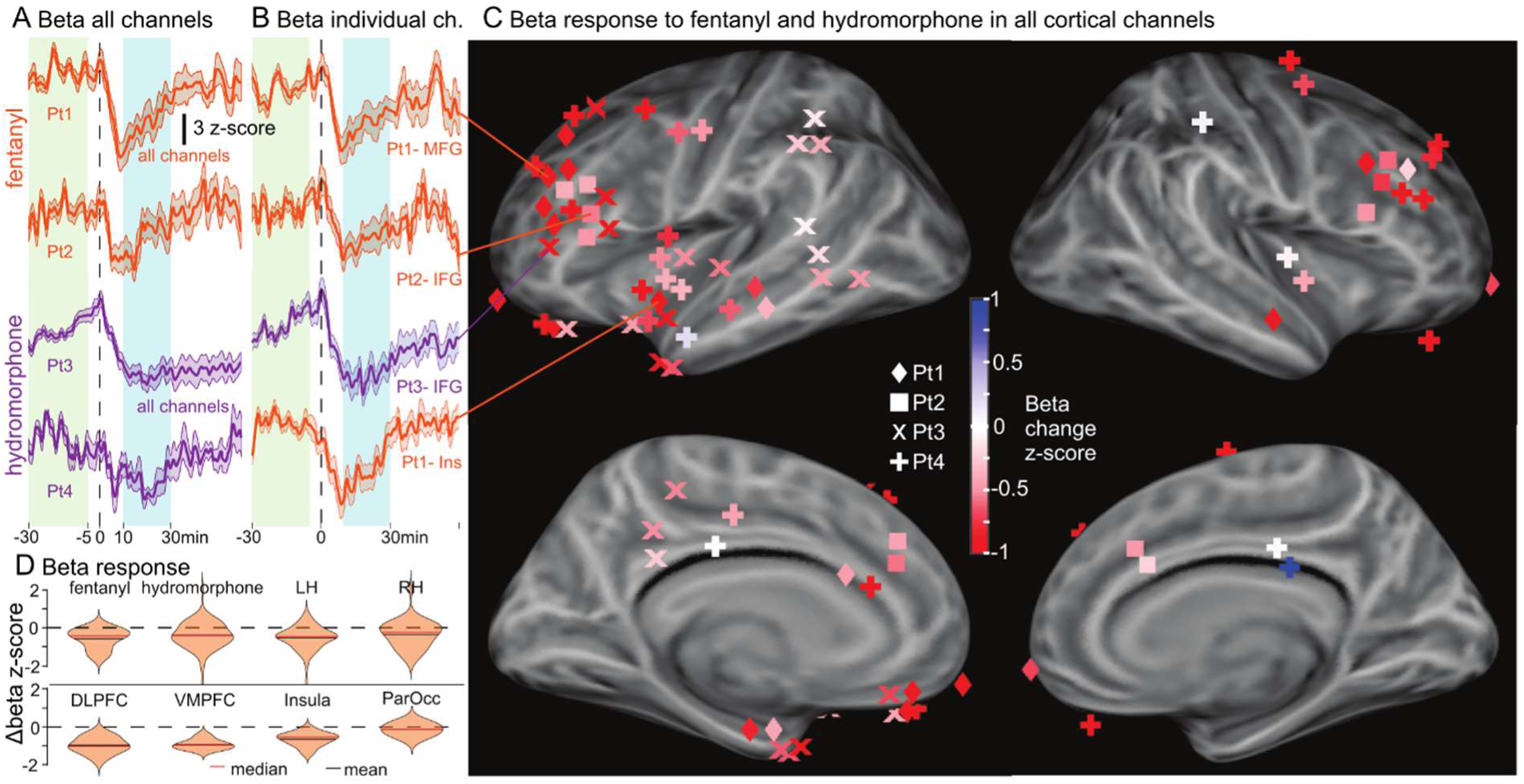
Beta amplitude modulation by intravenous opioids. **A.** Mean (±SEM) beta amplitude over trials and channels, aligned to opioid administrations. Post-drug beta suppression (cyan) is contrasted with baseline (green). Orange = fentanyl; purple = hydromorphone. All traces p<0.05 FDR corrected. **B.** Same as A, but for example sites. **C.** Beta modulation at all sites, color-coded. Opioids generally tended to reduce beta amplitude. Suppression was strongest in VPFC, DLPFC, Insular cortex. See Table 3 for significance testing. **D.** Distributions of site-wise modulation for various categories. Fentanyl and hydromorphone suppressed beta amplitude similarly; left hemisphere sites showed somewhat greater suppression.

**Table 3.** Linear mixed-effects model coefficients for evoked drug effects and pain-relief correlations in beta and high-gamma amplitude. Negative values (red) indicate that decreases in amplitude were related to drug effect or pain relief. Significant coefficients are bolded. All subject-wise random effects converged to zero under ML. R^2^ beta-drug = 0.49 ordinary, 0.43 adjusted; R^2^ HG-drug = 0.30, 0.22; R^2^ beta-pain = 0.24, 0.15; R^2^ HG-pain = 0.28, 0.19.

| Beta |  |  |  |  |
| --- | --- | --- | --- | --- |
| Model Term | Drug (z) | Pain (R) | p Drug | p Pain |
| Main Effect | **-0.47** | **-0.10** | 7.1E-14 | 0.01 |
| DLPFC | **-0.47** | 0.00 | 6.9E-06 | 0.97 |
| VPFC | **-0.38** | **-0.24** | 0.02 | 3.7E-02 |
| Parietooccipital | 0.23 | 0.10 | 0.24 | 0.47 |
| Insular | -0.09 | -0.12 | 0.53 | 0.19 |
| Lat. Temporal | 0.04 | -0.13 | 0.78 | 0.17 |
| Cingulate | 0.18 | -0.14 | 0.13 | 0.11 |
| Uncus | -0.14 | -0.03 | 0.53 | 0.86 |
| Lat. Amygdala | **-0.86** | 0.11 | 8.4E-07 | 0.32 |
| Med. Amygdala | **0.33** | **0.38** | 0.04 | 5.6E-04 |
| Anterior HC | **0.61** | **0.25** | 1.4E-04 | 0.02 |
| Posterior HC | **0.85** | 0.03 | 1.5E-07 | 0.77 |
| Hemisphere (LH) | **-0.13** | 0.01 | 0.01 | 0.78 |
| Drug Type (HMOR) | 0.02 | -0.05 | 0.69 | 0.16 |
| High Gamma |  |  |  |  |
| Model Term | Drug (z) | Pain (R) | p Drug | p Pain |
| Main Effect | -0.01 | -0.03 | 0.86 | 0.40 |
| DLPFC | 0.14 | 0.06 | 0.39 | 0.38 |
| VPFC | -0.25 | **-0.24** | 0.35 | 0.05 |
| Parietooccipital | -0.07 | -0.04 | 0.82 | 0.76 |
| Insular | **-0.49** | **-0.32** | 0.03 | 2.0E-03 |
| Lat. Temporal | -0.25 | 0.09 | 0.24 | 0.38 |
| Cingulate | **0.42** | 0.11 | 0.03 | 0.21 |
| Uncus | -0.27 | 0.13 | 0.44 | 0.40 |
| Lat. Amygdala | **-0.55** | 0.11 | 0.04 | 0.37 |
| Med. Amygdala | -0.18 | **0.36** | 0.48 | 2.4E-03 |
| Anterior HC | **0.58** | 0.17 | 0.02 | 0.14 |
| Posterior HC | **0.67** | -0.07 | 5.7E-03 | 0.54 |
| Hemisphere (LH) | **-0.23** | 0.25 | 0.01 | 0.52 |
| Drug Type (HMOR) | **0.28** | **-0.09** | 5.9E-03 | 0.02 |
| Key | drug n.s. - | pain n.s. - | drug n.s. | pain n.s. |
|  | **drug sig. +** | pain n.s. + | drug sig. | pain n.s. |
|  | **drug sig. -** | **pain sig. -** | drug sig. | pain sig. |
|  | **z = 1** | **R = 0.5** | drug sig. | pain sig. |
|  | **z = -1** | **R = -0.5** | drug sig. | pain sig. |

### Opioid effects on beta in amygdala and hippocampus

Distinct patterns emerged within the amygdala in response to opioids. In P1, a single depth electrode spanning the medial-lateral axis revealed a functional division: medial amygdala (MAmyg) exhibited a transient beta *increase* from 5-10 minutes post-FENT, while in lateral amygdala (LAmyg) FENT robustly *decreased* beta amplitude over ∼30 minutes (Fig. 4A,B). Beta responses to opioids in the hippocampus were variable, but tended to be small increases rather than decreases (Fig. 4C). Relative to the whole-brain effect, MAmyg and both anterior and posterior hippocampus (AHC, PHC) showed significantly greater beta amplitude post-opioid (Table 3).

**Figure 4.**
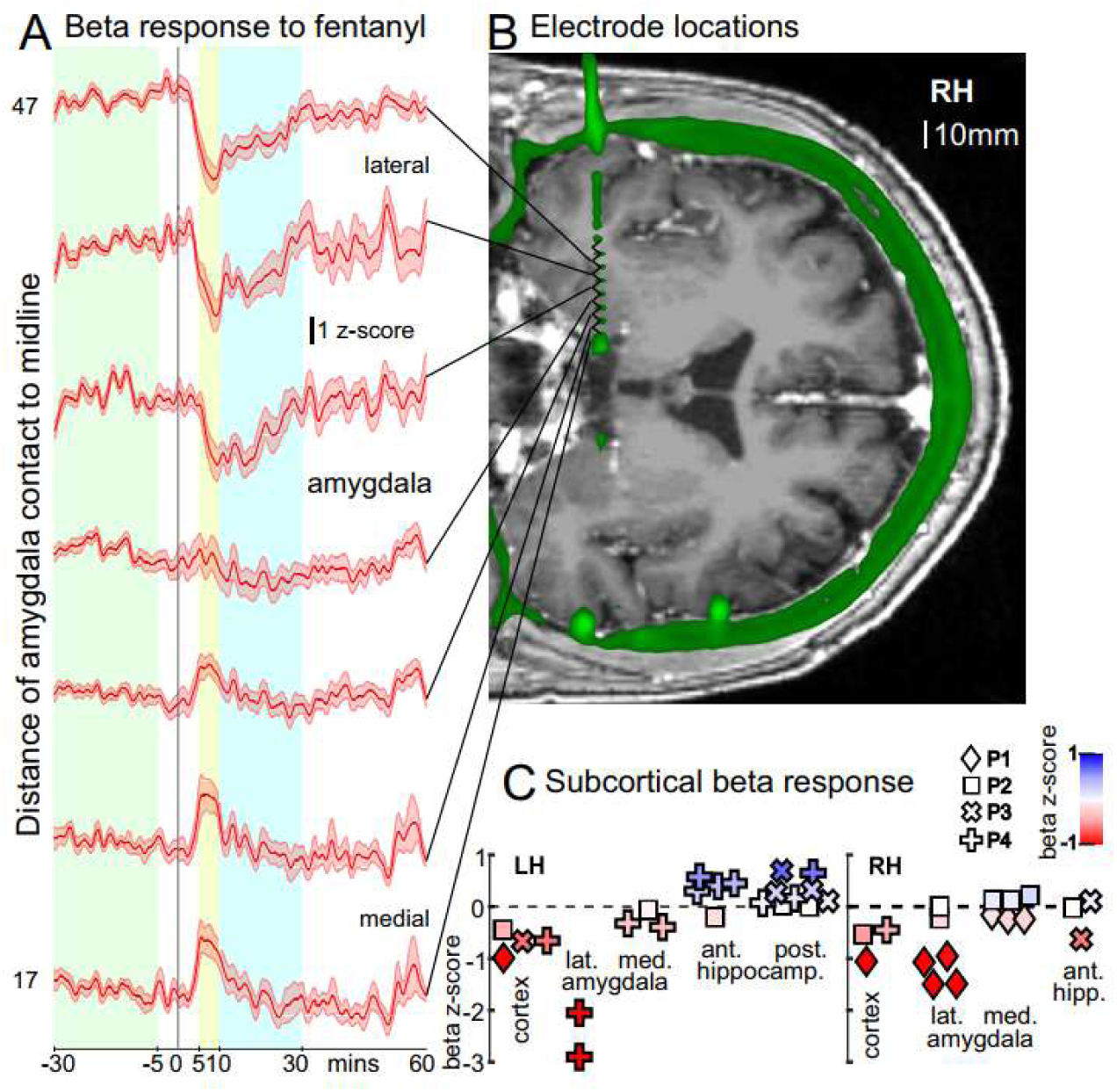
Beta modulation in medial temporal lobe. **A.** Beta amplitude aligned to opioid injection at sites along the medial-lateral trajectory of a single electrode spanning right amygdala. Lateral amygdala exhibited suppression (similar to cortex), while medial amygdala showed transient increases ∼5-10 minutes post-drug (p=0.004, 0.005, 0.027, FDR corrected, post-hoc). **B.** Coregistered MRI/CT showing locations of sites (bipolar pairs) represented in A. **C.** All sites in medial temporal structures. Lateral amygdala resembled cortex with beta suppression; medial amygdala responses were variable. Hippocampal responses were generally positive or null.

### High-gamma amplitude modulation by opioids

In contrast to the broad suppression of beta, high-gamma (HG) amplitude showed no overall trend following opioid administration (Fig. 2, Table 3). However, distinct site- and region-specific patterns emerged. HG amplitude was suppressed in a subset of cortical regions, including Insula (INS) and LAmyg, while other medial temporal regions (MAmyg, AHC, PHC) tended to show increases (Table 3, Fig. 5A,B). In cingulate cortex, HG tended to increase following opioids, driven largely by a subset of sites in mid-to-posterior cingulate. Finally, FENT was associated with lower HG amplitude than HMOR (Fig. 5C), in contrast to the similar beta suppression produced by both drugs.

**Figure 5.**
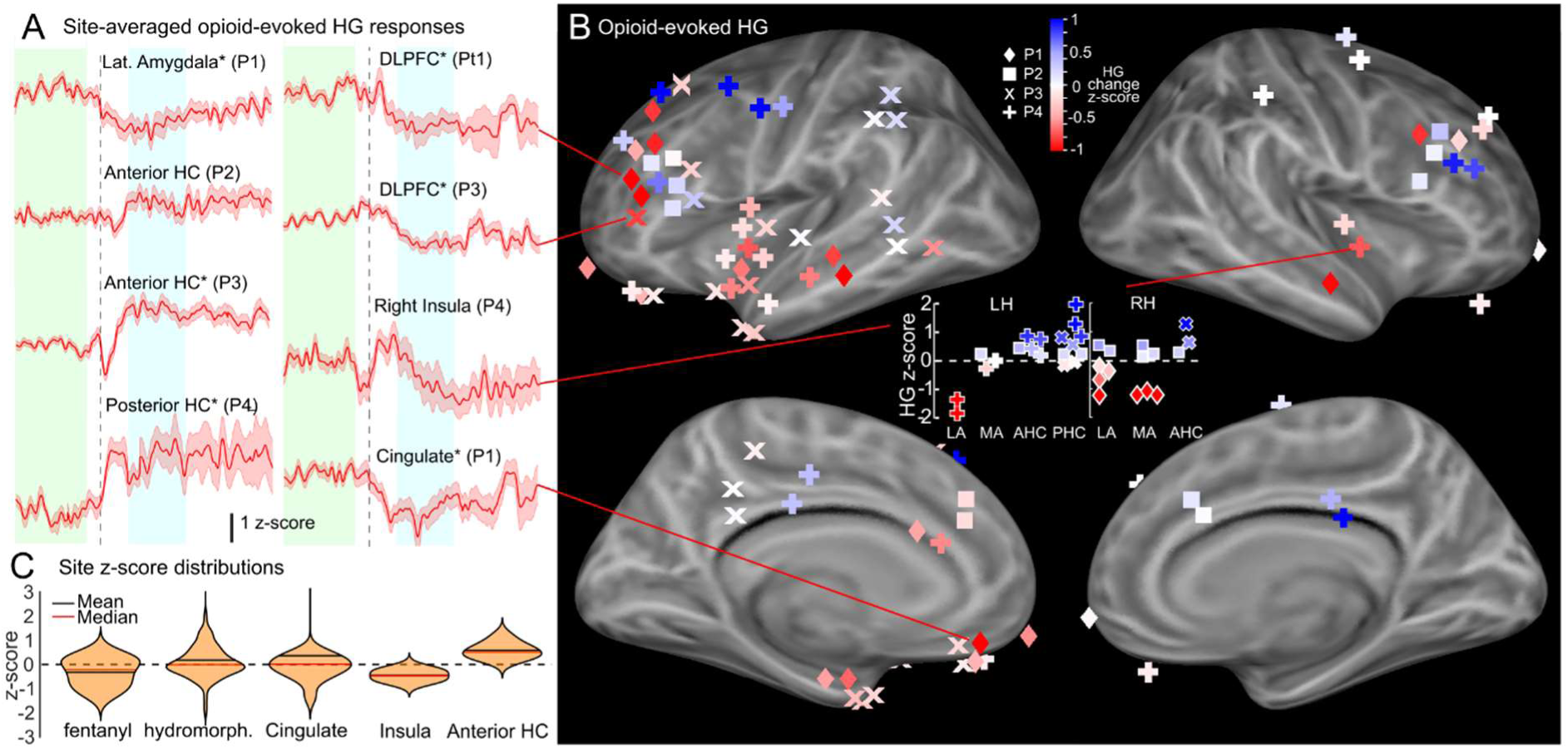
High-gamma amplitude modulation by intravenous opioids. **A.** Time-course of high-gamma amplitude at example sites. Evoked effects varied in direction and time-course. * Indicates significance of the individual site (p<0.05 FDR over all sites). **B.** Site-wise HG modulation, color coded. HG was suppressed in Insula and Lateral Amygdala, and increased in Cingulate, AHC, and PHC. See Table 3 for significance testing. **C.** Distributions of site-wise modulation by category. HG increases and decreases occurred in similar proportions.

### Neural correlates of opioid analgesia

To isolate neural correlates of pain relief specifically, we tested for correlations between beta or HG amplitude and degree of reported pain relief (change in VAS score pre-to-post drug), while holding opioid dosage constant. Amplitude changes were measured from a pre-drug baseline (30-10 min prior to administration) to the 10 minutes preceding the post-drug pain assessment. For each site, the correlation (R value) between neurophysiological change and pain relief was computed. We then modeled these R values as a function of drug-effect z-scores, and including random effects by subject to account for heterogeneity. Both beta (β=0.12, p=0.0025) and HG (β=0.08, p=0.049) significantly predicted pain relief across sites (Table S1), indicating that opioid modulation of these signals tracks the signature of pain relief.

Notably, this effect was driven by strong contributions from specific regions. Suppression of beta in VPFC and of HG in INS correlated with pain relief. Conversely, in the medial temporal lobe, pain relief was beta (MAmyg and AHC) or HG (MAmyg only) *increases* (Fig. 6, Table 3).

**Figure 6.**
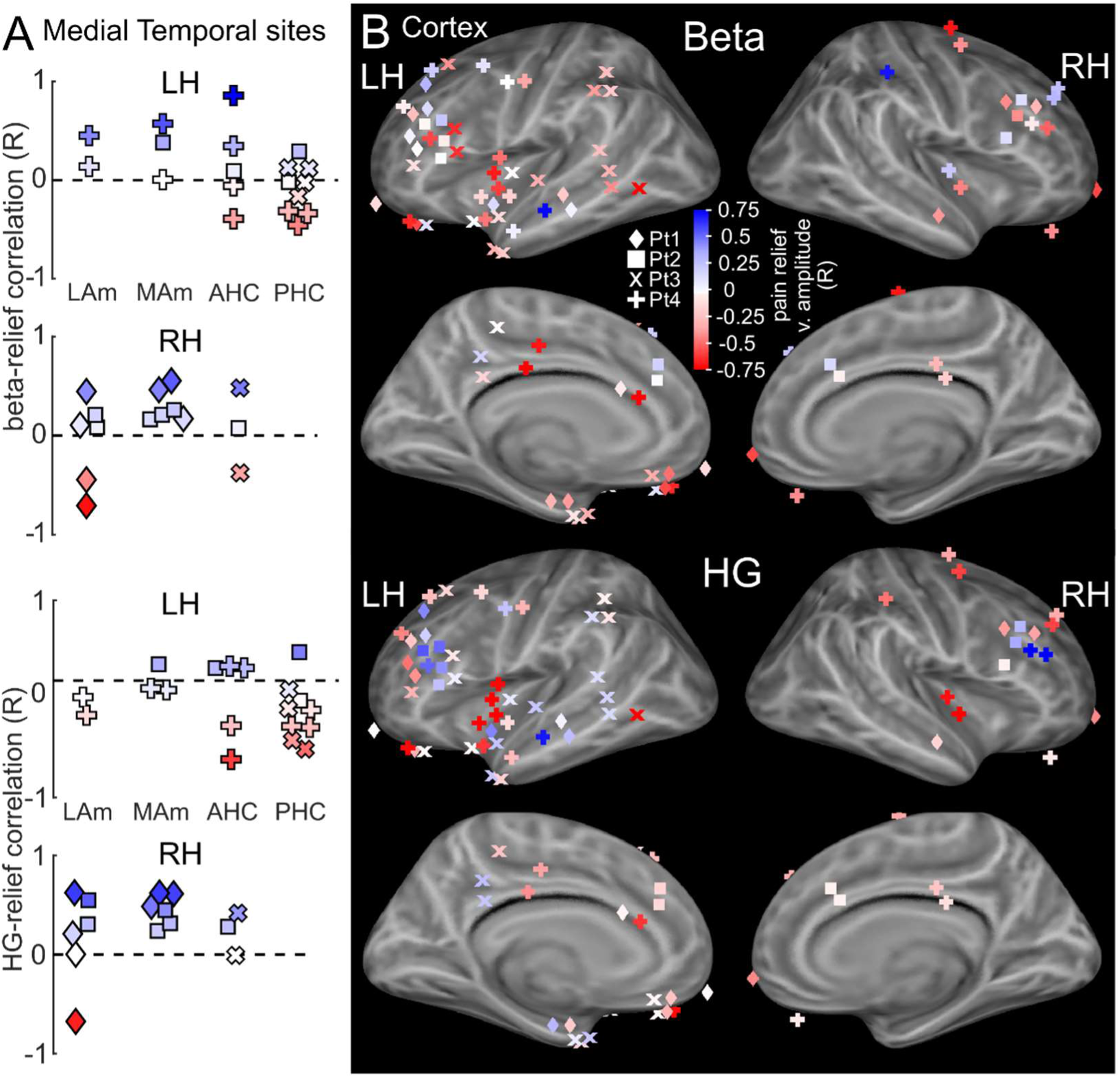
Site-wise correlations between amplitude changes and pain relief. **A.** Correlation (R-values) between pain relief (VAS pre-to-post drug) and beta (top) or HG (bottom) amplitude change in medial temporal sites. **B.** As in A, for cortical sites. See Table 3 for significance testing.

### Controls for nurse interaction and trial heterogeneity

In general, opioids were administered in a subset periodic structured nurse visits, thus there was a corresponding set of nurse visits in which no opioid was given. To verify that neurophysiological changes were not the result of social interaction, pain evaluation, or the interruption of ongoing activities, we analyzed and tested these control non-opioid nurse interactions, applying procedures identical to those used to evaluate opioid effects. None of the region-wise or overall effects reported above to be observed in relation to opioid administration were also evoked by nurse check-ins (Table S2).

Additionally, to account for trial-to-trial variability in clinical context and signal quality, we fit a linear mixed-effects model with random intercepts by trial and subject. All opioid-evoked effects reported above were robust to this control (Table S3).

## Discussion

The present study represents the first characterization of the neurophysiological effects of opioids using intracranial EEG in humans, and is among the first efforts to isolate the neural correlates of pain relief in EEG generally [1,41,64]. Our dataset represents a sample of clinically-relevant pain, which may provide a more naturalistic and nuanced pathophysiological model than the more commonly studied transient, experimenter-evoked painful stimuli [19,44]. Though preliminary, these findings provide novel insight into the therapeutic effects of opioids beyond early sensory systems, on substrates of interpretive and affective processes responsible for the psychological impact of pain.

The μ-opioid receptor is densely expressed in forebrain structures including the amygdala, anterior hippocampus, anterior cingulate cortex, orbitofrontal cortex, and insula [42]. PET rCBF and BOLD fMRI studies have generally reported signal increases following opioid administration, most consistently in anterior cingulate, but also involving caudate, thalamus, and amygdala [1,41,45,72]. In contrast, FDG-PET revealed decreases in many of the same structures [47], suggesting that blood flow-sensitive measures may be confounded by direct vascular effects of opioids [59,76]. A few studies have combined acute pain stimuli with opioid administration to attempt to isolate analgesia-related signals [1,41]. Notably, Oertel et al. [64] observed a dose-dependent attenuation of pain-evoked BOLD signal in insula, while a separate cluster, centered more anteriorly in ventral prefrontal cortex, was step-wise suppressed by moderate alfentanil doses. They interpreted this as a dissociation between sensory (insula) and affective (VPFC) components of pain, a model closely aligned with the present findings (see below).

Intracranial evoked potentials and high-frequency activity following acute experimental pain have generally observed responses in somatosensory cortex, insula, anterior cingulate, orbitofrontal cortex, and amygdala, consistent with known telencephalic pain pathways [8,9,12,23,28,46]. Recently, two iEEG studies have utilized machine learning trained on neurophysiological features [77] (and in one case, on facial expressions [39]) to predict spontaneous pain state. In both cases, the authors trained within-subject models to predict spontaneous high/low pain state transitions. These approaches can yield powerful individualized models (with Huang et al. in particular reporting impressive classifier performance); however, they do not permit statistical inference about shared predictive features. In contrast, our cross-subject framework isolates generalizable predictors of opioidergic pain relief with direct translational relevance.

No prior studies have modeled pain state across participants by shared EEG/LFP features, which may be hampered by confounds underlying spontaneous variations in pain as well as high subject heterogeneity. To circumvent these effects, we utilized the baseline period prior to rapidly-acting opioid administration as a control to isolate predictors of pain relief.

We observed that opioids induced a widespread reduction in beta-frequency amplitude, most prominent in the prefrontal cortex, as well as suppression of high-gamma amplitude in the insula. Effects in medial temporal lobe were mixed, with suppression of beta and HG in lateral amygdala, but relative increases in medial amygdala (beta) and hippocampus (beta and high-gamma). We compared beta and HG amplitude changes between equal opioid doses yielding variable pain relief to determine which effects specifically predicted relief. These included beta decreases in VPFC, HG suppression in insula, and beta increases in medial amygdala and anterior hippocampus. Furthermore, there was a significant tendency over all sites to positive correlation between opioid effects and the association of pain relief with beta and HG, further supporting the interpretation that these neural processes play a broad role in opioidergic pain relief. Our findings are consistent with prior work implicating these regions, and the beta frequency band in particular, in clinical pain [30,90].

Pain-specific responses, dissociated from neutral sensory stimulation and proportional to intensity [16], have been observed in both anterior [38] and posterior insula [28,51], in studies utilizing iEEG and fMRI. In cortical models of pain, the posterior insula is often considered the central hub for sensory processing of painful stimuli [30,66,75], while anterior insula is associated with pain salience [53,86]. Consistent with this, we observed a tendency for opioids to suppress high-gamma amplitude in this region, along with a correlation between HG suppression and pain relief. Our sampling did not allow separate analysis of anterior and posterior insula. Given the critical role of insula in pain processing, future work should further resolve opioid-evoked dynamics in this region, especially interactions with other forebrain regions subserving attention and affect.

In ventral prefrontal cortex (VPFC), we found that beta oscillations, rather than high-gamma, were associated with opioidergic pain relief. Beta activity is linked to active attention, top-down control [5,73], and maintenance of the current cognitive and sensorimotor state [25]. A recent large-scale iEEG study identified beta as the most widespread oscillation in sensorimotor, frontal, and limbic regions, particularly in reinforcement-linked networks [13]. In scalp EEG, beta oscillations are enhanced in chronic pain conditions, especially over frontal regions [4,20,79,82,90]. Thus, beta oscillations represent a promising biomarker candidate at the intersection of pain sensation, aversion, and opioid action. Among executive regions, the VPFC is particularly implicated in affective evaluation [18,70,74]. Spontaneous fluctuations in chronic pain are associated with medial and ventral PFC BOLD activation [6,7,31]. VPFC lesions alter the influence of expectation on pain perception [55], and VPFC activation represents the relative valuation of acute painful stimuli [88]. Based on the present findings, we speculate that suppression of beta-band activity in VPFC may interrupt the ‘capture’ of evaluative attention by pain, diminishing its affective impact and perceived severity. This interpretation aligns with models emphasizing the interpretive and affective dimensions of clinical pain [34,48,87].

Recent research implicates the hippocampus in pain processing and the development of inflammatory or chronic pain states [33,84]. Chronic pain is associated with reduced hippocampal gray matter volume [57,63], while increased hippocampal volume has been observed following non-pharmaceutical pain treatment [29]. μ-opioid activation increases the excitability of the hippocampus by disinhibiting pyramidal neurons [52] and releasing astrocytic glutamate [60]. We observed an increases in both beta and high-gamma amplitude in anterior and posterior hippocampus, but only anterior beta was evoked by opioids and correlated with pain relief. Beta may be more prominent in the human hippocampal LFP than in model species [37,54]. and increased beta coherence between hippocampus and amygdala occurs during fear memory retrieval [85].

Similarly, we observed that opioids increased relative beta power in medial amygdala, and that this increase was associated with pain relief. Rapidly evoked beta was observed in medial amygdala sites (Fig. 4A), distinct from suppression in adjacent lateral sites. As a central node integrating limbic processing and physiological responses, the central amygdala (CeA) sends prominent antinociceptive descending projections to the periaqueductal gray [68], and regulates pain according to behavioral state [10,35,36,61,62]. In rats, CeA lesion or inactivation attenuates morphine analgesia in the tail-flick assay [49], suggesting a conserved antinociceptive role.

While medial temporal beta has not been directly linked to pain relief, beta oscillations may occur particularly in dopamine-modulated networks [13], and opioids strongly influence subcortical limbic structures via activation of the ventral tegmental area and consequent dopamine release [40,80,81]. Opioid-evoked dopamine release occurs with a very short latency, consistent with the rapid beta onset we observed (Fig. 4) [89]. A plausible mechanism is that dopamine receptor activation in MAmyg and AHC, perhaps in concert with MOR activation, jointly engages descending antinociceptive pathways. Accordingly, intra-VTA morphine is analgesic in a rat model [3]. This provides a neural substrate linking salience, appetitive drive, and reinforcement (dopamine) to antinociception (MAmyg) on a context-sensitive basis (AHC), implementing reward-induced (and, in other contexts, stress-induced) analgesia.

Due to limited sampling, we could not characterize regional interactions; future work utilizing prospective designs will be essential to clarify the mechanisms of beta modulation in opioid analgesia. Additionally, opioids were administered in an naturalistic clinical environment, where external factors may have influenced neural activity or subjective analgesia. Findings should therefore be generalized cautiously, though results were robust to multiple controls for confounds and heterogeneity (Tables S2,S3).

Overall, four out of six limbic structures, but none of the five non-limbic regions, showed significant changes in beta and/or high gamma related to pain relief (Fig. 7). These results suggest a model in which opioid analgesia engages distinct limbic mechanisms: (1) nociceptive gating (amygdala, anterior hippocampus); (2) decreased concerned fixation on pain (VPFC beta suppression); and (3) dampened pain sensation (insula HG suppression). Though preliminary, these results may inform future investigation into the pathophysiology of clinical pain and targets for novel analgesic therapies.

**Figure 7.**
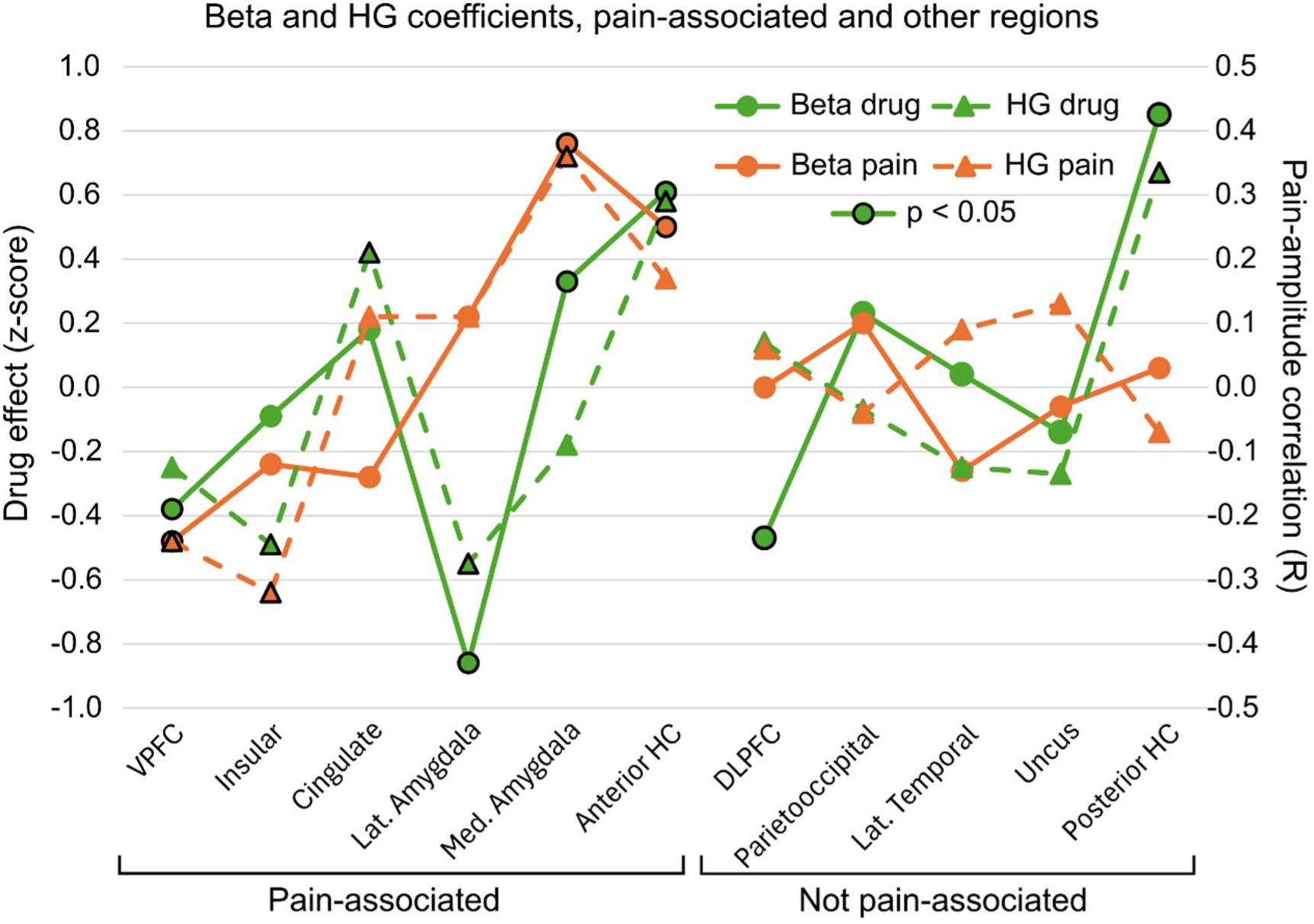
Pain-relevant regions show drug-pain correlations. Pain relief correlations (orange; beta=solid/circles, HG=dashed/triangles) were found only in classically pain- or affect-related regions (VPFC, Insula, Medial Amygdala, Anterior HC). In these, drug-evoked changes (green; beta=solid/circles, HG=dashed/triangles) were predictive of relief. No correlations were found in regions not associated with pain.

## Data Availability

All analysis code will be made available via GitHub. sEEG data, relative opioid administration and control event times, cortical reconstructions/electrode localization will be available on ieeg.org.

## Data Availability

All analysis code will be made available via GitHub. sEEG data, relative opioid administration and control event times, cortical reconstructions/electrode localization will be available on ieeg.org. See published version for links to data and code repositories.

## Acknowledgements

The authors express their sincere thanks to Syd Cash, Burke Rosen, Ilya Verzhbinsky, and Adam Niese for their contributions to this work.

