## Supplementary Figure and Tables for "Opioidergic pain relief in humans is mediated by beta and high-gamma modulation in limbic regions"

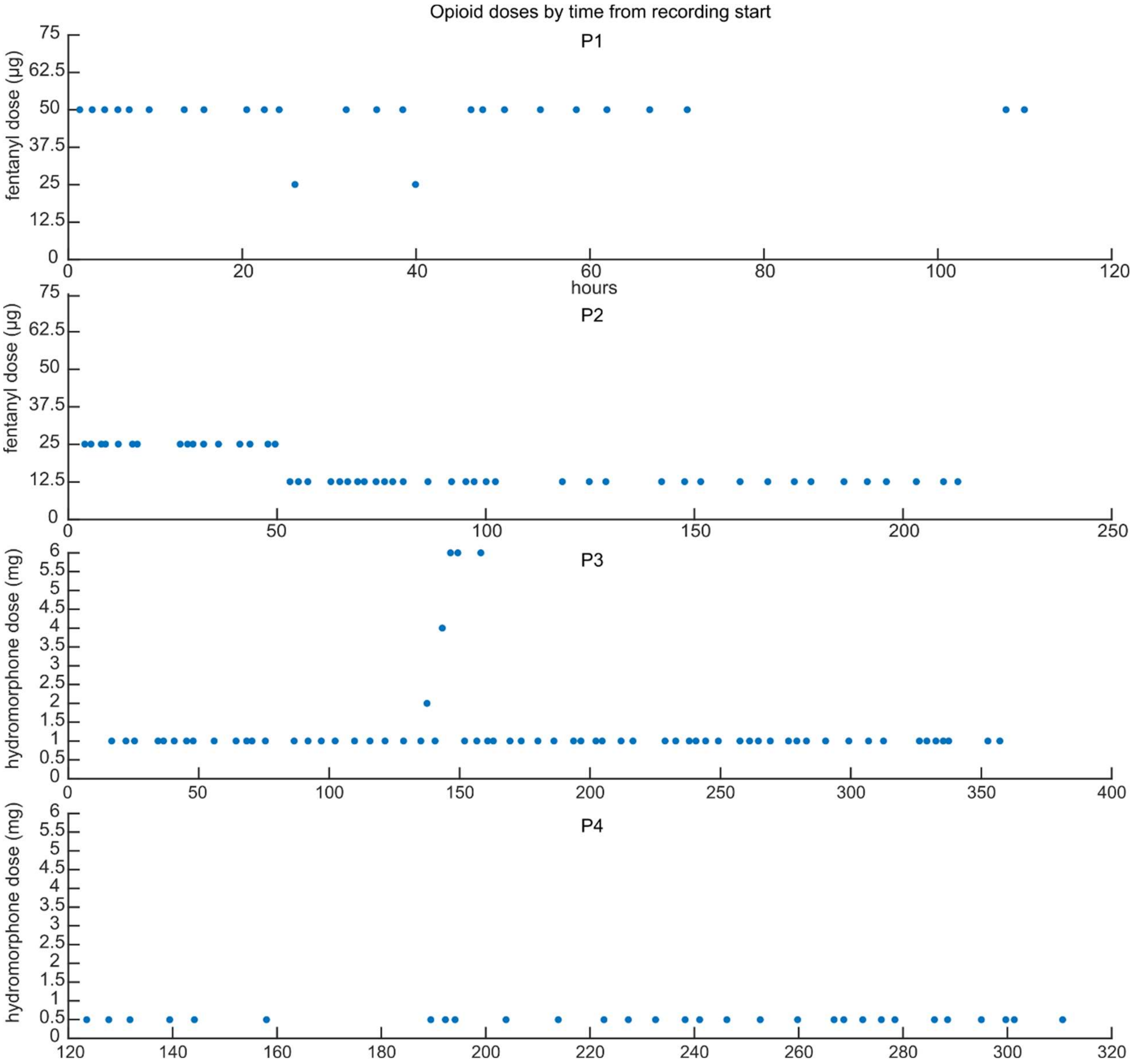


**Figure S1. Timing and dosage of IV opioids for all participants.** Only doses included in analyses of drug effects are shown. Participants P1 and P2 received fentanyl; P3 and P4 received hydromorphone. N doses: P1=22, P2=33, P3=51, P4=14. All doses were spaced at least one hour apart.

### Supplementary Tables

| | z-score | Intercept | Subject (SD) |
|---|---|---|---|
| Beta | **0.12** | -0.05 | 0 |
| p | 0.03 | 0.28 | — |
| HG | **0.08** | 3E-3 | 0.17 |
| p | 0.05 | 0.97 | — |

**Table S1. Specificity of drug effect for pain-relief correlates.** Mixed-effects linear models assessed whether opioid-evoked changes tended to track the neural signature of pain relief. Amplitude-pain-relief correlations (R value) were modeled as a function of drug effect (z-score), accounting for inter-subject variance. Results show a positive and significant relationship for both beta amplitude (0.12 R/z, adjusted $R^2$=0.03) and high gamma (0.08 R/z, adjusted $R^2$=0.17).

| Evoked Coeff. (z-score) | No opioid | | Opioid | |
|---|---|---|---|---|
| Model Term | Beta | HG | Beta | HG |
| Main Effect | 0.10 | **0.20** | **-0.45** | -0.02 |
| DLPFC | -0.01 | 0.01 | **-0.46** | 0.13 |
| VPFC | -0.03 | 0.04 | **-0.38** | -0.23 |
| Parietooccipital | -0.03 | -0.18 | 0.28 | -0.03 |
| Insular | -0.13 | -0.13 | -0.11 | **-0.50** |
| Lat. Temporal | -0.03 | **0.21** | 0.05 | -0.25 |
| Cingulate | **-0.25** | **-0.36** | 0.18 | **0.42** |
| Uncus | 0.22 | -0.20 | -0.13 | -0.31 |
| Lat. Amygdala | **0.27** | **0.32** | **-0.86** | **-0.54** |
| Med. Amygdala | 0.14 | 0.01 | **0.32** | -0.18 |
| Anterior HC | -0.05 | 0.16 | **0.60** | **0.53** |
| Posterior HC | -0.14 | 0.02 | **0.85** | **0.72** |
| Hemisphere (LH) | 0.04 | -0.03 | **-0.13** | **-0.23** |
| Subject (STD) | 0.09 | 0.17 | 0 | 0 |
| p-value | No opioid | | IV opioid | |
| Model Term | Beta | HG | Beta | HG |
| Main Effect | 0.05 | 0.03 | 3.1E-13 | 0.82 |
| DLPFC | 0.81 | 0.93 | 9.9E-06 | 0.39 |
| VPFC | 0.74 | 0.74 | 0.03 | 0.37 |
| Parietooccipital | 0.73 | 0.21 | 0.17 | 0.92 |
| Insular | 0.07 | 0.21 | 0.42 | 0.02 |
| Lat. Temporal | 0.70 | 0.04 | 0.69 | 0.24 |
| Cingulate | 7.3E-05 | 1.3E-04 | 0.16 | 0.03 |
| Uncus | 0.05 | 0.23 | 0.55 | 0.37 |
| Lat. Amygdala | 1.4E-03 | 0.01 | 1.1E-06 | 0.04 |
| Med. Amygdala | 0.07 | 0.96 | 0.05 | 0.47 |
| Anterior HC | 0.54 | 0.16 | 2.5E-04 | 0.03 |
| Posterior HC | 0.06 | 0.84 | 1.8E-07 | 3.0E-03 |
| Hemisphere (LH) | 0.18 | 0.48 | 0.01 | 0.01 |
| Key | negative, not sig. | | not significant | |
| | **negative, significant** | | significant ($p<0.05$) | |

**Table S2. Linear mixed-effects model coefficients for control (non-opioid nurse check-in) vs. opioid responses.** Models assess evoked beta and HG amplitudes during control (non-opioid) events and changes following opioids; these models are presented side-by-side for comparison. (Note that "Opioid" corresponds to the "Drug" columns in Table 3.) Negative coefficients (red) indicate suppression. Bolded values are statistically significant. No significant effects in control events matched the direction of drug-evoked responses. $R^2$ (conditional) beta-control = 0.36 ordinary, 0.28 adjusted; $R^2$ HG-control = 0.35, 0.27; $R^2$ beta-drug = 0.49, 0.43; $R^2$ HG-drug = 0.30, 0.22.

| Fixed Effects | Beta | | High Gamma | |
|---|---|---|---|---|
| | Coeff. (z) | p | Coeff. (z) | p |
| Main Effect | **-0.47** | 1.6E-06 | -0.01 | 0.97 |
| DLPFC | **-0.50** | 4.1E-19 | 0.06 | 0.49 |
| VPFC | **-0.21** | 0.02 | -0.17 | 0.19 |
| Parietooccipital | **0.23** | 0.01 | 0.11 | 0.41 |
| Insular | -0.11 | 0.20 | **-0.45** | 3.7E-04 |
| Lat. Temporal | **0.16** | 0.02 | 0.00 | 0.97 |
| Cingulate | 0.04 | 0.53 | 0.12 | 0.23 |
| Uncus | -0.10 | 0.26 | -0.15 | 0.27 |
| Lat. Amygdala | **-0.60** | 3.2E-08 | **-0.36** | 0.02 |
| Med. Amygdala | **0.30** | 1.3E-03 | -0.18 | 0.18 |
| Anterior HC | **0.34** | 5.3E-05 | **0.52** | 2.5E-05 |
| Posterior HC | **0.81** | 7.3E-27 | **0.39** | 3.6E-04 |
| Hemisphere (LH) | **-0.09** | 4.8E-03 | **-0.18** | 1.6E-04 |
| Random Effects | STD | | STD | |
| Trial | 0.61 | | 0.71 | |
| Subject | 0.14 | | 0.34 | |

**Table S3. Linear mixed-effects model of opioid responses by trial.** Random effects included for subjects and trials. All findings observed in site-level analyses remain significant when controlling for trial-level heterogeneity. Conditional $R^2$ beta-drug = 0.24 ordinary, 0.24 adjusted; $R^2$ HG-drug = 0.16, 0.15

| Region of Interest | HCP-MMP1 parcels |
|---|---|
| Dorsolateral Prefrontal Cortex (DLPFC) | SFL, 6v, 8BM, 8Av, 8Ad, 9m, 8BL, 9p, 8C, 44, 45, 6r, IFJa, IFJp, IFSp, IFSa, p9-46v, 46, a9-46v, 9-46d, 9a, i6-8, s6-8 |
| Ventral Prefrontal Cortex (VPFC) | 10r, 47m, 10d, 47l, a47r, 10v, a10p, 10pp, 11l, 13l, OFC, 47s, pOFC, p10p, p47r |
| Parietoocippital Cortex | V1, V6, V2, V3, V4, V8, V3A, POS2, V7, IPS1, FFC, V3B, LO1, LO2, PIT, PCV, 7Pm, 7m, POS1, 5m, 5mv, 5L, 7AL, 7Am, 7PL, 7PC, LIPv, VIP, MIP, LIPd, PFt, AIP, ProS, TE2p, TPOJ3, DVT, PGp, IP2, IP1, IP0, PFop, PF, PFm, PGi, PGs, V6A, VMV1, VMV3, V4t, V3CD, LO3, VMV2, VVC |
| Insular Cortex | OP4, OP1, OP2-3, 52, PFcm, PoI2, FOP4, MI, AVI, AAIC, FOP3, FOP2, PoI1, Ig, FOP5, PI |
| Lateral Temporal Cortex | MST, MT, A1, PSL, STV, RI, TA2, STGa, PBelt, A5, STSda, STSdp, STSvp, TE1a, TE1p, TE2a, TF, PHT, PH, TPOJ1, TPOJ2, FST, MBelt, LBelt, A4, STSva, TE1m, ITG |
| Cingulate Cortex | RSC, 23d, v23ab, d23ab, 31pv, 23c, 24dd, 24dv, p24pr, 33pr, a24pr, p32pr, a24, d32, p32, 31pd, 31a, 25, s32, a32pr, p24 |
| Uncus | Pir, EC, PreS, H, PeEc, PHA1, PHA3, TGd, PHA2, TGv |
| Lateral Amygdala | Basolateral, Lateral Nuclei (manual localization) |
| Medial Amygdala | Corticomedial, Central, and Basomedial Nuclei (manual localization) |
| Anterior Hippocampus | Hippocampal head (manual localization) |
| Posterior Hippocampus | Hippocampal body, tail (manual localization) |
| Rolandic | 4, 3b, FEF, PEF, 55b, SCEF, 6ma, 1, 2, 3a, 6d, 6mp, 6a, 43, FOP1 |

**Table S4. Specification of regions of interest.** Cortical ROIs were defined using HCP-MMP1 atlas parcels. Hippocampus and amygdala were manually localized using co-registered CT/MRI.

| | P1 | P2 | P3 | P4 |
|---|---|---|---|---|
| DLPFC | 7 | 7 | 3 | 8 |
| VPFC | 3 | 0 | 2 | 2 |
| Parietooccipital | 0 | 0 | 4 | 1 |
| Insular | 1 | 0 | 2 | 8 |
| Lat. Temporal | 5 | 0 | 5 | 2 |
| Cingulate | 2 | 4 | 3 | 5 |
| Uncus | 0 | 0 | 4 | 0 |
| Lat. Amygdala | 4 | 2 | 0 | 2 |
| Med. Amygdala | 3 | 4 | 0 | 2 |
| Anterior HC | 0 | 2 | 2 | 4 |
| Posterior HC | 0 | 2 | 4 | 3 |
| Rolandic | 0 | 0 | 0 | 4 |
| Total | 25 | 21 | 29 | 41 |

**Table S5. Number of recording sites by region and subject.**
